# Transnational adoptees in healthcare: barriers, resources, and needs

**DOI:** 10.1101/2024.05.01.24306620

**Authors:** Mattias Strand, Natte Hillerberg

## Abstract

**Background:** After decades of research output, it is well established that transnational adoptees—i.e., individuals who are placed for adoption outside their country of birth—exhibit an increased risk of various negative mental health outcomes. Even so, there is a lack of suggestions for preventive measures or treatment interventions targeting the transnational adoptee population in the literature.

**Objective:** To explore experiences, opinions, and needs among adult transnational adoptees in Sweden concerning healthcare in general and mental healthcare in particular.

**Methods:** Sixty-six adult transnational adoptees residing in Sweden, born in 15 different non-European countries, were recruited for individual in-depth interviews about their experiences and opinions regarding psychosocial support and healthcare. The interview data were analyzed employing a codebook thematic analysis approach.

**Results:** Three overarching themes were identified: a) barriers to adequate treatment, b) helpful resources in dealing with health-related issues, and c) health-related needs and suggestions for the development of adequate support. Identified barriers include a lack of insight into and interest in adoptee health, colorblindness and unwillingness to address racism, expectations of gratitude, steep financial costs, lack of support from adoptive parents, and mistrust of support structures that involve adoptive parents or adoption organizations. Participants also describe helpful resources, such as the community of fellow transnational adoptees. Health-related needs and suggestions include more well-defined and easily accessible structures of support, improved knowledge and competence, a broader psychotherapeutic repertoire that better addresses adoption-related themes, improved support in situations that can be particularly stressful for adoptees (such as during pregnancy and as new parents), routine follow-up during childhood and adolescence, and education targeting adoptive parents. The need for greater attention to the well-being of children of transnational adoptees is also highlighted.

**Implications:** Based on these findings, a number of recommendations can be made. For example, knowledge about adoptee health should be strengthened, and psychotherapeutic competence in addressing issues related to racism should become a priority. After over 20 years of discussion, one or more national research and knowledge hubs on transnational adoption should be created. Moreover, economic resources should be made available to support transnational adoptees in accessing adequate treatment.

## 1 Introduction

It has been estimated that over 1 million children have been adopted across national borders since the end of World War II (1). Domestic adoption of children whose biological/first parents are deceased or cannot provide for them has been practiced in human societies since ancient times. In contrast, transnational adoption—i.e., when children are placed for adoption outside their country of birth—is a more recent phenomenon, emerging after World War II and evolving on a broader scale in the aftermath of the Korean War (2). From the 1960s and onwards, transnational adoption became the dominant form of adoption in many parts of the Global North, as fewer children were put up for domestic adoption in these countries (3). Unlike in earlier decades, however, a surplus of prospective adoptive parents currently exists due to socioeconomic development and altered policies in many origin countries. As countries such as South Korea and India have seen the emergence of large middle-class populations, the legitimacy of sending children abroad for adoption has been questioned (4,5) and accompanied by a parallel move toward domestic adoption in these countries (2,3). Moreover, countries such as Ethiopia and Guatemala have banned adoption to foreign countries altogether after reports of illicit activities and trafficking of children (6,7).

Research on the mental health of transnational adoptees has revealed alarming tendencies on a group level. Compared to the general population, transnational adoptees exhibit an increased risk of psychotic disorders (8), substance use (9,10), disordered eating (11,12), suicide attempts, and suicide (10,13,14) and are more likely to be in specialist outpatient and inpatient psychiatric treatment (10,15,16). Also, they display more symptoms of attention-deficit/hyperactivity-disorder (17), and externalizing behavioral problems (16,18,19) that might require residential care during adolescence (20). Moreover, transnational adoptees are more often unemployed and dependent on social welfare, are less likely to be in a relationship and to have children, and when they do have children they are more likely to be single parents (15,21). These findings are even more striking considering that a majority of transnational adoptees have typically been raised in middle- or high-income adoptive families, who tend to experience overall better health outcomes than the larger population (10,22).

Numerous biological, environmental, and societal factors have been suggested to explain the observed differences in mental health between transnational adoptees and non-adoptees; a more detailed account is beyond the scope of this article. As a general tendency, it can be noted that a previous research focus on pre-adoption factors such as adverse environmental impact during pregnancy (17) or maltreatment and neglect at children’s institutions (3,16) has more recently been supplemented by a critical view on the contributing role of post-adoption factors such as racism and ‘colorblindness’ that may leave transnational adoptees having to manage racialized societal stereotypes on their own without much support from their adoptive families or the society at large (23–30). Even though the exact mechanisms are not fully understood, the increased risk of negative mental health outcomes among transnational adoptees must now—after decades of research output on the topic—be considered an established fact (31). The relative lack of suggestions for preventive measures or treatment interventions targeting the transnational adoptee population in the literature is therefore somewhat surprising. It is reasonable to assume that this lack of suggestions in the medical field on how to improve the mental health of transnational adoptees can contribute to a prevailing and ultimately harmful notion of adoptees as ‘psychologically damaged’ by default (32,33).

There are currently approximately 60 000 transnational adoptees in the Swedish population (31,34). As a country of 10 million inhabitants, Sweden thus has the largest per capita proportion of transnational adoptees in the world. Adoptions to Sweden peaked in the 1970s and early 1980s and have declined substantially since 2005 (35), which means that a majority of transnational adoptees in the country are now adults. Sweden has a relatively long history of epidemiological research on the health of transnational adoptees (31); this work has contributed to raising awareness but has resulted in few public health implementations targeting the group. Based on available evidence, an official report of the Swedish government, published in 2003, recommended the introduction of mandatory parental education classes for adoptive parents, strengthening competence on adoption-related issues across the lifespan among healthcare professionals, and the establishment of a national research and knowledge hub on transnational adoption, modeled on existing services in the Netherlands (36). As far as we can tell, none of these recommendations have been implemented on a national level.

### 1.1 Aim

The overall aim of this study was to explore experiences, opinions, and needs among adult transnational adoptees in Sweden concerning healthcare in general and mental healthcare in particular. The two main research questions were:

i. What experiences of seeking and receiving mental healthcare exist among transnational adoptees? This included aspects such as available resources and support in society, barriers to care, experiences of addressing the adoption background in therapy, the importance of racism and racialization for healthcare, etc.
ii. What can and should be done to improve the mental health of transnational adoptees? This included ideas on how to organize and provide healthcare, preferred therapist characteristics, changing needs throughout the life course, etc.

## 2 Materials and methods

### 2.1 Study design

For this study, a qualitative research design based on individual in-depth interviews was used. A preliminary semi-structured thematic interview guide was developed based on findings from previous studies on the non-healthcare experiences of transnational adoptees. A reference group of six individuals, all transnational adoptees who were either representatives of one of the organizations for transnational adoptees, involved in research on adoption, or therapists with experience working clinically with the group, was created. The reference group provided feedback on the preliminary interview guide, and adjustments were made accordingly. The study design also allowed for iterative fine-tuning of the interview guide; for example, when the needs of children of adoptees emerged as an important topic in the initial interviews, this was included as a potential interview topic when relevant.

The explicit focus of this study was on the experiences of transnational adoptees; therefore, national or domestic adoptees were not targeted for inclusion. Although the experiences of national adoptees are, of course, important in their own right, they are not necessarily readily comparable with those of transnational adoptees. For example, transnational adoptees typically have very little available information about their early lives, whereas this is often not the case for national adoptees. Moreover, most Swedish national adoptees are White and do not face racism and discrimination of the kind that tends to affect transracial adoptees (3,18).

### 2.2 Participants

To be eligible for inclusion in the study, participants had to be ≥18 years of age and have a history of being transnationally adopted. Participants were recruited by disseminating information about the study on websites, in newsletters, and on social media accounts of various Swedish organizations for transnational adoptees (such as Adopterade etiopiers och eritreaners förening, Adopterade koreaners förening, Chileadoption.se, Organisationen för vuxna adopterade och fosterbarn, Svenska koreaadopterades nätverk and Transnationellt adopterades riksorganisation). These networks have previously been used to recruit study participants in qualitative research on topics other than health and healthcare (37,38). Interviews were conducted with 65 adult participants. One additional participant wished to provide written answers instead of partaking in a face-to-face interview; the total number of participants was thus 66. Eight participants were male, and 58 were female. Among those 61 participants who mentioned their age, the mean age was 43.4 years (median: 43, range: 21-58 years). In terms of adoption background, the 65 participants whose countries of birth were known were adopted from 15 different countries in East Africa; South, Southeast, or East Asia; South America; and Oceania. The most common country of birth was South Korea (47.7%), followed by India (10.8%), Ethiopia (7.7%), Chile, Colombia, and Sri Lanka (all 6.2%), reflecting the characteristics of the adult Swedish adoptee population (35).

### 2.3 Procedures

Individual in-depth interviews lasting between 30 and 120 minutes were performed over the course of 11 months. NH performed 14 and MS performed 51 of the interviews. All interviews were digitally recorded and transcribed verbatim. The transcripts were pseudonymized by omitting or altering potentially identifying details, such as names, place of residence, and occupation; also, when participants were born in a country that has sent relatively few adoptees to Sweden, the name of the country was altered.

### 2.4 Analysis

The transcribed interview data were analyzed employing what is sometimes referred to as a codebook thematic analysis approach (39), combining the methodological framework outlined by Braun and Clarke (40) with elements of conventional qualitative content analysis (41). Moreover, due to the protracted interview procedure, preliminary stepwise analyses of the collected data were performed as suggested by Malterud in her account of systematic text condensation (42), eliciting a gradually developing understanding and allowing for sharpening of the focus along the way. In choosing this framework, the aim of the analysis was to describe important themes in the participant narratives rather than to theorize or speculate. In a highly iterative process, the data was first read and re-read and initial ideas for coding categories and overarching themes were drafted. Second, the transcripts were coded and labelled according to a ‘bottom-up’ principle, avoiding as far as possible preconceived ideas about adoptee experiences and needs. Third, the categories were grouped into themes. No predefined criteria were applied in determining what would constitute a separate theme; instead, meaningful clusters were developed inductively by analyzing recurrent patterns in the interview data. Fourth, the themes were reviewed and reworked collaboratively using mapping techniques to achieve a reasonable structure for describing the data. Finally, illustrative pseudonymized quotes were chosen and the findings were presented and contextualized.

### 2.5 Reflexivity

Importantly, the descriptive approach described above does not automatically involve the assumption that themes somehow pre-exist in or “emerge from” the data; reflexivity and interpretation were still important aspects of the research process. During the interview phase of the study, both authors continuously reflected on their own positions in relation to the participants, the study context, and the research questions. These reflections were recorded in the form of “field” notes. An obvious aspect that may have affected the interview situation is that both authors are White non-adoptee clinician-researchers interviewing adoptees mostly identifying as persons of color about experiences relating to transnational adoption, “in-betweenness”, and racism. This dynamic was usually explicitly addressed at the beginning of the interview; for example, even when a participant had no specific questions or queries before starting the interview, the interviewer would explain that s/he her/himself was neither an adoptee nor an adoptive parent. A typical participant response would then be that they were used to this or that it did not matter much. Several participants responded that it was good to know that no adoptive parents were involved in the study. One participant said that s/he felt that it would have been easier to talk to a fellow adoptee or a person of color; however, reflecting on the findings reported below regarding therapist identity, it is reasonable to assume that more participants experienced this as a problem, although they did perhaps not feel comfortable addressing it. On the other hand, participants occasionally stated that they felt it was easier to talk to a non-adoptee about their experiences. Much effort was put into identifying and counteracting barriers for participants to feel safe and to be able to provide a full narrative. This involved learning from feedback from the reference group on contextual framing and establishing a non-rigid interview procedure. However, based on the findings reported below—involving clear preferences for adoptee therapists or therapists of color among many of the participants—it can be assumed that a race-of-interviewer effect (43) might have impacted on participants’ willingness and ability to talk openly about their experiences.

### 2.6 Ethics and preregistration

This study was conducted in accordance with the ethical standards of the Helsinki Declaration of 1975, as revised in 2008. The study was approved by the Swedish Ethical Review Authority (Nos. 2022-03422-01 and 2023-03465-02). Written consent was obtained from all participants. The study protocol has been preregistered on the Open Science Framework (osf.io/256ns).

## 3 Results

Three overarching themes were identified in the interview data: a) barriers to adequate treatment for transnational adoptees, b) resources that are helpful in dealing with health-related issues, and c) health-related needs and suggestions for the development of adequate support. For each of these themes, several subthemes were identified (see Figure 1 and Table 1); these subthemes are described in more detail below.

**Figure 1.**
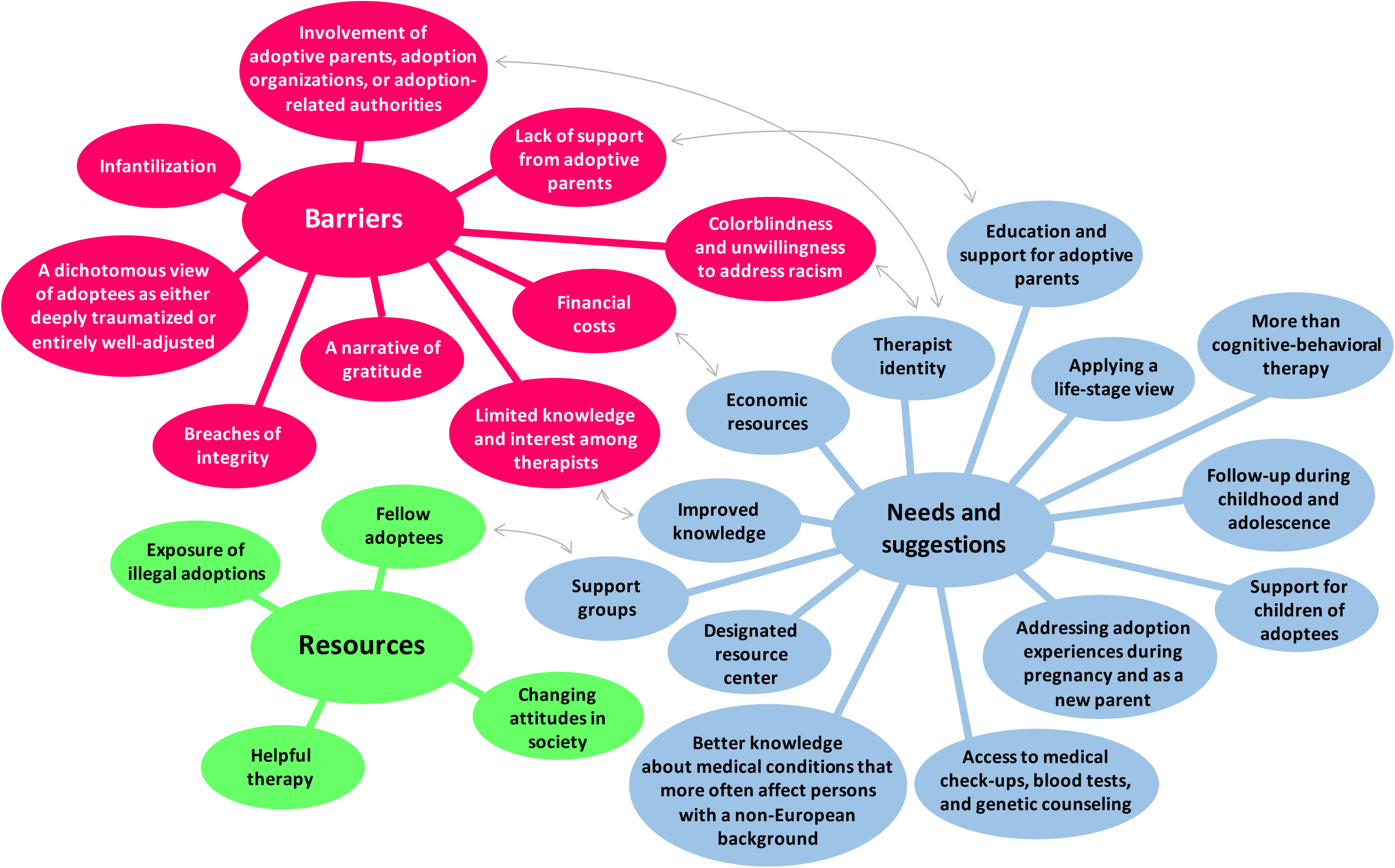
Themes and subthemes. The arrows represent particularly strong relationships within the data.

**Table 1.**
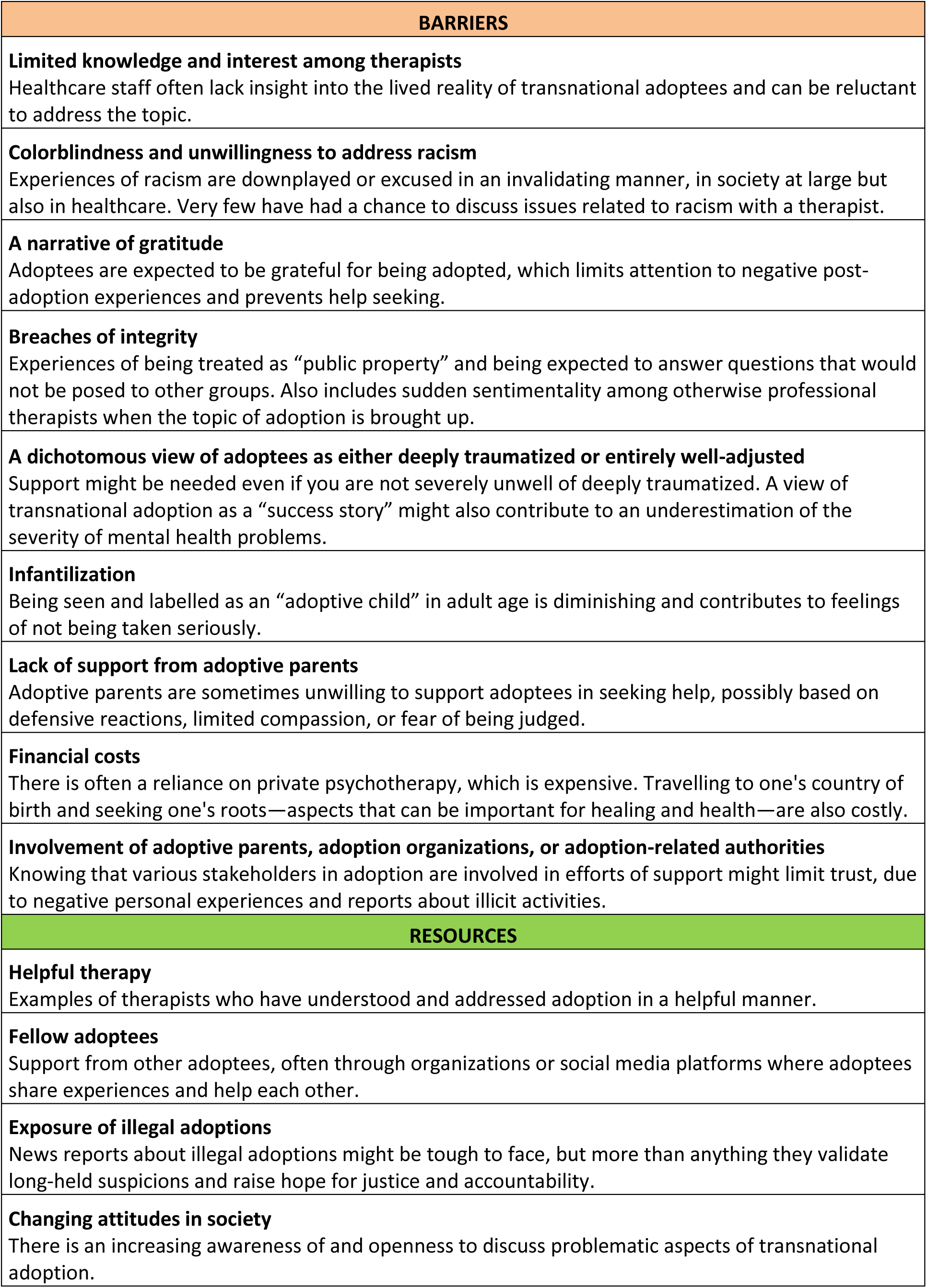
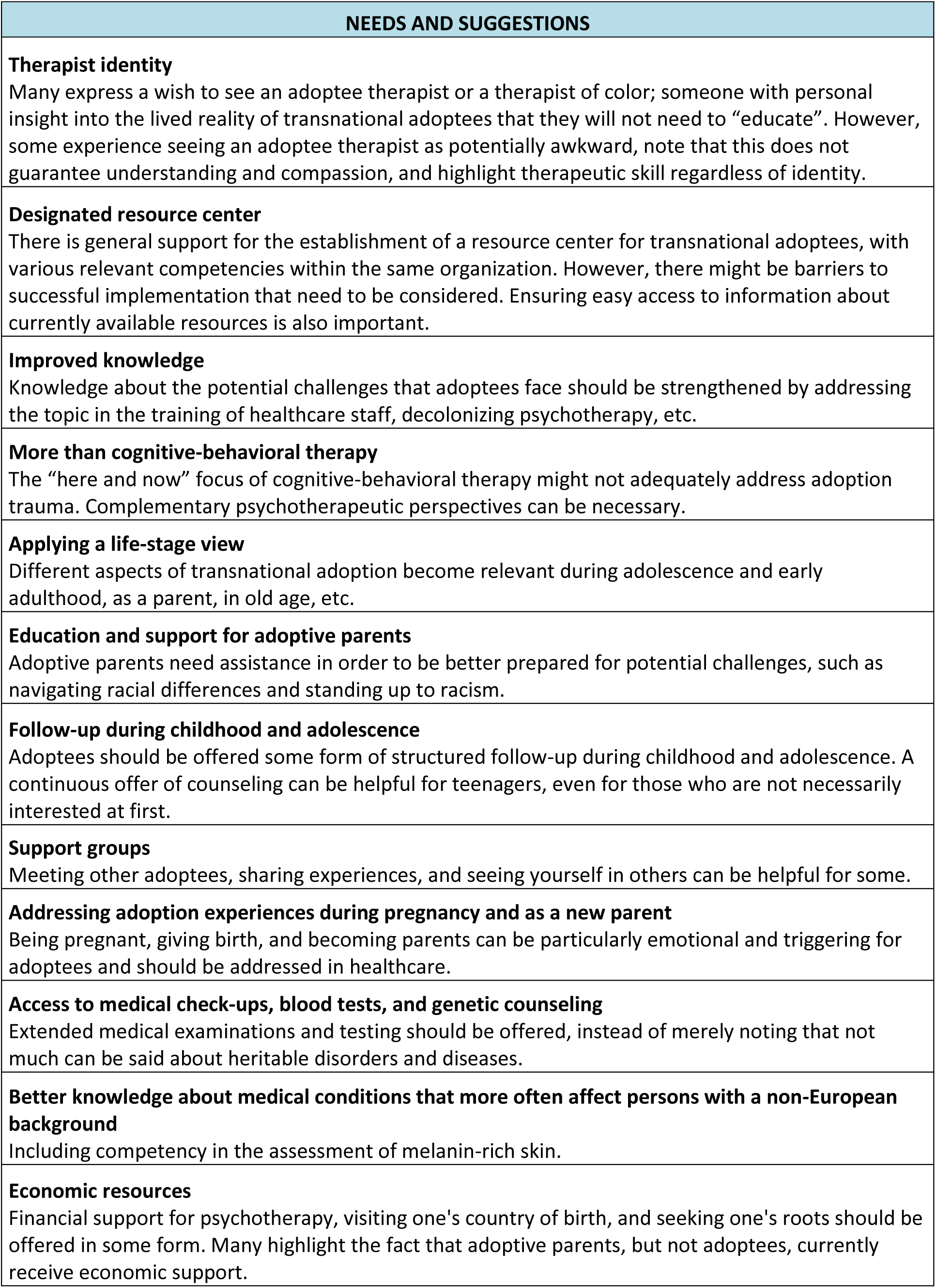
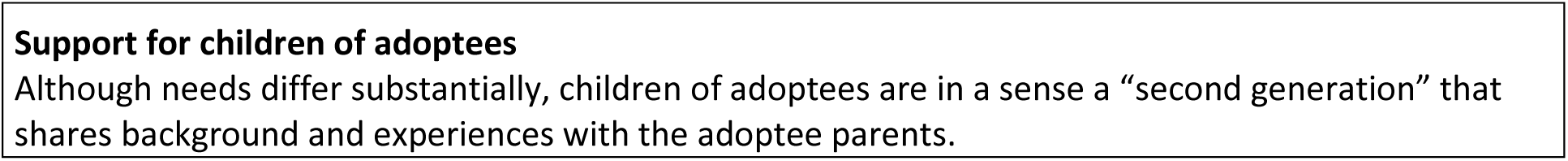
Themes and subthemes, with brief descriptions.

### 3.1 Barriers

#### 3.1.1 Limited knowledge among therapists

Most participants experience limited knowledge about adoption-related issues among therapists and other healthcare professionals as a major barrier in accessing adequate therapeutic support. This can take the form of poor insight into factors relevant to adoptee health:

> *It’s stressful to realize that the person who’s supposed to help you has a much more limited understanding of attachment, trauma, and adoption than you yourself. It’s not the adoptee’s job to teach the therapist.*
>
> *It was as if everyone understood that “she’s an adopted girl and that has probably left a mark on her somehow”, but nobody wanted to talk about it because they didn’t know how to. And those who did address it sometimes did it in a very awkward and ignorant way.*
>
> *Psychotherapists have occasionally commented on the fact that I’m adopted, but then haven’t been able to elaborate on why that might be significant.*

More often, however, participants describe a persistent reluctance among therapists to even address the lived experiences of transnational adoptees:

> *I really want to stress that I’ve never been asked what it’s like to be adopted. It hasn’t ever been raised in any contact I’ve ever had in healthcare.*
>
> *No one asked me explicitly, and it doesn’t feel like it’s been there implicitly, beneath the surface, either. It’s as if my life started here in Sweden.*
>
> *I mean, I might have mentioned that “oh right, I’m adopted”, but no one really picked up on it or asked questions about that specifically. It’s been more like “so how was your childhood?”, without making that connection. I’ve thought about that. It’s been hard for me to push it on my own. Because somehow, I’ve sought help so that someone can assist me in putting these pieces together.*

#### 3.1.2 Colorblindness and unwillingness to address racism

In addition to a reluctance to address adoption-related issues more generally, many participants describe a specific unwillingness among therapists to discuss experiences of racism and how they may impact health:

> *Therapists must understand all these pieces that adoptees… It’s not just about attachment, because they’re well versed in that area as a part of their psychologist training. But racism, minority stress [i.e., stress faced by members of minority groups due to stigma and discrimination]—there are all these other things that they need to relate to or deal with.*
>
> *There was never any discussion about how I could handle it, it was more like I just shouldn’t be bothered about it. And that [the offenders] didn’t know any better. So, I feel that I… I wasn’t given any tools to deal with it and there was also very little emotional support.*
>
> *And this last time, I had to try to explain what it’s like to be brown and to experience hostility and to be afraid just because of the color of my skin. I’ve been beaten by skinheads just for… When I talk about being afraid because of the color of my skin, it’s a real thing.*

Some participants see this as a manifestation of a longstanding exceptionalist Swedish self-image as a ‘post-racial’ society:

> *I suppose it’s also connected to how we don’t like to talk about race in Sweden, how we would rather be colorblind and think that this race talk only adds to racism. While, to understand and raise awareness about these structural issues, we actually need to address them. […T]he elephant in the room is race.*

This type of casual colorblindness in healthcare is brought up as insulting and harmful by several participants:

> *I think they’re trying to tell me that I’m worth just as much as any White Swede. And I’m thinking: well, sure. But there’s a risk that they invalidate my specific experiences of being racialized, and…*
>
> *Healthcare staff need to be more confident about what is and what isn’t racism. Acknowledging that you’re from another ethnic background and taking that into account isn’t racist. On the contrary, noting that you’re from another ethnic background and ignoring it—that’s racism. And I think most people are like: “Well, at least I wasn’t racist because I didn’t bring it up”. While in this case, that’s actually the racist part.*
>
> *They say that “no, I think of you as Swedish”, but that’s also kind of weird because you wouldn’t say that to any other Swede, would you?*

#### 3.1.3 A narrative of gratitude

A prevailing societal ‘narrative of gratitude’—the notion that transnational adoptees ought to feel thankful for having been “rescued” into adoption and that any critique of adoptive parents or of adoption on a structural level is ungrateful and unwarranted—is described as a major barrier to healthcare by a large number of participants. In relation to health specifically, many experience that the mere existence of illness or the act of seeking help are implicitly seen as signs of ungratefulness. This notion has often become internalized, at least in part:

> *I even heard from my school nurse when I was little that I ought to [feel] fortunate instead. So that caused—what can I call it?—shame, I think. About seeking help as an adoptee. I think many of us feel… We feel like we don’t dare to say anything, because then we’re complaining.*
>
> *They’ve been the most wonderful [adoptive] parents. But I still don’t feel comfortable talking to them about everything, because I keep thinking that if I bring up the fact that I’m not doing so well, then I’m being ungrateful.*
>
> *There’s a fear of being a burden, expressing my needs, and saying that this has been difficult for me. Perhaps you’ve heard all your life that “yes, but there are others who are worse off” or “you would have died if you had…” I’ve heard that all the time: “Well, you wouldn’t have survived if you had remained in Ethiopia.” Lately, I’ve actually started saying: “But how can you be so sure about that?”*
>
> *I don’t believe in the idea that it’s all those things that happen before the adoption that traumatize you; it’s a lot about what happens after the adoption. Such a sequence of events—that anything post-adoption is at least better than what came before— underestimates the experiences here in Sweden and invalidates one’s feelings.*

#### 3.1.4 Breaches of integrity

Several participants describe how recurrent experiences of being expected to answer all types of intimate questions about their background have made them more hesitant to open up to others, including healthcare staff:

> *I’ve been asked [in a non-healthcare setting]: “Why did your parents adopt you? Couldn’t they have children of their own?” And I’m like… What makes you think that I’m talking to my parents about… Sometimes I’ve even given idiot answers—that they didn’t know how to make a baby. Just because I get so tired of it.*
>
> *Your boundaries are being violated in how they talk about you, how they talk about you in relation to your Swedish family, how they talk about the Chinese family or this whole process, the money that’s involved and all that. When you’re a child and you don’t have any answers at all… It’s an enormous disregard for boundaries.*
>
> *It’s like you’re an attraction that everyone just has to ride. That’s how it feels sometimes.*

Other examples of breaches of personal integrity include descriptions of sudden outbursts of sentimentality among otherwise professional therapists when the topic of adoption is brought up:

> *I had a great psychotherapist the last time I was in therapy, but then I mentioned something about having met my biological family. And then the perspective was suddenly: “Oh, that’s so exciting! Wow!” And I would rather she had said something like: “But hey, how’s that been for you?” I’ve had that several times. They become fascinated by the fact that I’ve met my biological family. And then it’s suddenly not about me.*
>
> *During our last session, I told him that I’d found my family and that I was going back. You know, to visit them some time. And then he started crying and told me that “you found your way home” or something like that. And I wasn’t really there myself yet.*

#### 3.1.5 A dichotomous view of adoptees as either deeply traumatized or entirely well-adjusted

A few participants mention that preconceived notions of adoptees as severely unwell or deeply traumatized have affected them negatively, as if they are somehow destined to suffer:

> *I learned early on that adoptees are overrepresented in psychiatry and when it comes to suicide. And that was like… Well alright, what should I do with that information? I’m 14.*

However, this also means that adoptees who present with less severe problems are often not seen as worthy of attention in healthcare:

> *I told them that I felt very lost in my thoughts on adoption and how it has affected my everyday life, and that it’s been hard for me to find myself in all of this. And then I was compared to others who suffered really bad. It was like they were comparing us: “I can see that you’ve had a rough time, but there are adoptees who’ve had a horrible childhood, who’ve been beaten, and they made it.”*

Participants also bear witness to a parallel view of transnational adoption as a “success story” that might contribute to an underestimation of the severity of mental health problems:

> *I’ve always been very adaptable and very high functioning, as seen from the outside. That’s why psychologists have often let me go, because I seem to be doing so well.*

These co-existing notions create a false dichotomy that leaves little room for nuances and fluctuations across the lifespan and ultimately makes it more difficult to access adequate support. Furthermore, participants describe how this binary also tends to be used to pit adoptees against each another, labeling them as “good” and “bad”, causing further harm to the adoptee community as it creates suspicion and distrust among peers who could otherwise rely on each other for help.

#### 3.1.6 Infantilization

A few participants describe a tendency among healthcare staff and others to not treat them as adults, based on their adoptee status:

> *We’re not being listened to; they don’t give us any attention. They don’t think we’re capable of speaking for ourselves. That we’re still children—adoptive children even as adults.*
>
> *And it’s so infantilizing to hear that “you’re an adoptive child”. Say what now? No, I’m an adult person who was adopted as a child. I’m a parent, and I’m entering middle age, and I’ll grow old here. It’s so reductive and diminishing to call us adoptive children. It really is, and it’s ignorant.*

#### 3.1.7 Lack of support from adoptive parents

Many participants talk about their Swedish adoptive parents as unsupportive when it comes to addressing mental health issues and seeking healthcare. Importantly, this tendency is observed both among those who describe a genuinely bad relationship with their adoptive parents and among those who describe an otherwise fairly unproblematic relationship:

> *They were born in the 1940s; they don’t talk about emotions*.
>
> *I was very young [the first time I went to the hospital], and I remember dad saying something along the lines of “we mustn’t tell anyone about this”. And that stuck with me deeply, so I never talked to anyone about it.*
>
> *I hinted to my parents that it might be a good idea for them to see a therapist, too. And then they—I don’t know what they said, but the point was like “we’re not lunatics”. They’re old school. They think you’re completely wacko if you seek [therapy].*
>
> *As an adoptive parent, some may perhaps feel that they need to be so much better. It’s my impression when I talk to my mother, this sense of failure when a child ends up in [mental] healthcare. And on top of that, they were academics and highly educated and high earners. For them, it just didn’t add up. It was challenging for them to accept help and look at their own role. It was easier to say, “but Jenny’s the one who’s acting up, it’s Jenny who cuts herself, she needs to go to a million therapy sessions”. But the fact that they were a part of it, they couldn’t really—they wouldn’t acknowledge that.*
>
> *Especially when my family had to be dragged along to the appointments, I thought that was awful.*

#### 3.1.8 Financial costs

Many of the participants describe that they have had to rely on private psychotherapy to work on various adoption-related issues, which rapidly becomes very costly. Furthermore, traveling back to reconnect with one’s country of birth or searching one’s roots through archival requests at adoption facilities, DNA tests, etc.—activities that many participants see as closely connected to their psychological well-being—are also associated with steep financial costs:

> *I mean sure, if you wish to spend [400 USD] every month to be in treatment with a private therapist, you can do that. But it’s a hell of a lot of money if you’re doing it for a year or two. It’s an unreasonable sum. And I can also feel that—I feel like what the hell… I’m not the one who decided I should come here.*
>
> *All the [therapists] who specialize in adoptees are private, so it gets pretty expensive. And I felt that it was holding me back, because I kept thinking: “Oh shit, this’ll cost me [120 USD] an hour, I’d better be good at it. I have to deliver and get as much out of it as possible.” I mean, the fact that it was so expensive distracted me.*

#### 3.1.9 Involvement of adoptive parents, adoption organizations, or adoption-related authorities

Finally, many participants mention that knowing that various stakeholders in adoption are involved in efforts of support may severely limit trust, due to negative personal experiences and/or reports about illicit activities related to adoption:

> *I believe that the element of trust is really, really important. You have to feel safe that [the therapist] isn’t pushing their own agenda; that the support is unconditional.*
>
> *It would definitely be problematic if I saw a psychologist who wasn’t transparent about [being an adoptive parent], I mean from the start. And if they were, I would probably see it as a major issue and ask to see someone else.*
>
> *And then I see the [Swedish adoption organization] logo underneath, and I feel like “no, wait, that’s for people who want to adopt”. And it sounds kind of horrible, but I feel that they’re not on my side. They’re a bit biased, they’re still mostly there for the parents. […] And I haven’t been proven wrong unfortunately, in feeling that way—it’s mor4e like the impression has grown stronger over the years because of their own statements.*
>
> *It doesn’t feel like the Swedish authorities care that much about us yet. The same goes for this [support structure launched by the government]: it’s more like something they started because all these scandals were brought to light.*

### 3.2 Resources

#### 3.2.1 Helpful therapy

Several participants mention previous experiences of helpful psychotherapy and other treatment contacts in mental healthcare. These therapist contacts are characterized by higher levels of knowledge about adoption-related issues, but perhaps even more so by an openness to address adoption as an important element in a patient’s life, regardless of the level of specific expertise:

> *In my recent psychological assessment, [the psychologist] was very open to these conversations, and knowledgeable—she had studied the material.*
>
> *She was very robust, down to earth, and she wasn’t afraid of—how should I say?—she wasn’t afraid of darkness or whatever you want to call it. The inner fears and despair of another human. Well, all this stuff that exists and hurts so bad.*

#### 3.2.2 Fellow adoptees

Discussing one’s experiences with fellow adoptees in Sweden through, for example, adoptee organizations or on online platforms is repeatedly mentioned as a helpful resource in terms of psychological well-being:

> *The only safe spaces I can think of are with others, in communities: in various Facebook groups or in organizations, among other adoptees. It’s the only context in which I feel understood, and like I understand others.*
>
> *When others talk about their problems, I also realize: “Oh my god, those are things that happen to me all the time. And this is actually racist, it’s actually crazy that people… Yes, I also have this attachment issue. Oh my god.”*
>
> *They [i.e., organization for Korean adoptees in Sweden] are so immensely important. It’s difficult to imagine what we would’ve done or where we would’ve been without them. That would’ve been… And it also goes to show that for a really long time, there’s been this enormous need, and we created a community and a way to show support.*
>
> *It was rewarding just being around people who look like me, who have similar experiences. It was a personal thing. I didn’t have a goal or a purpose, it just felt good to be in that context.*

#### 3.2.3 Exposure of illegal adoptions

A few participants describe the recent reports in Swedish and international media on the widespread occurrence of illegal adoptions as anxiety-provoking. However, most of the participants that talk about the subject actually see the media exposure in a more positive light and describe it as a recognition of sorts that may ultimately make it easier for adoptees to address negative experiences:

> *It’s a good thing actually. I’ve felt enormously validated by finally seeing something other than this happy story that everybody seems to want to talk about and emphasize.*
>
> *It’s been really satisfying for me that this debate has been raised. I feel like I don’t have to explain myself to everyone. And perhaps I dare to hope that more people get to understand what it’s like to be an adoptee. So that I don’t have to fight this battle on my own.*
>
> *These are problems that I’ve known about for a few years before it caught media attention and so on. So, I think it’s very important and it’s very good. And hopefully it might lead to further investigation that can—my wishful thinking—make things better for us adoptees. It might contribute to more and better resources for us. Or just knowledge and awareness.*

#### 3.2.4 Changing attitudes in society

Some participants reflect upon how various types of changing attitudes in society have been beneficial for them as adoptees. These changes are seen as reflecting a decreasing homogeneity of the Swedish population in terms of race and ethnicity, greater awareness of adoption-related issues, and less stigmatizing views on psychological distress and help seeking:

> *I guess that since Sweden has become more multicultural, you don’t look so different anymore. You might say it makes life easier.*
>
> *The therapists I’ve had are more up-to-date now. Compared to 2004, when my therapist told me that “nooo, I think chinky eyes are super pretty” […]. And nowadays they’ll say: “Yes, I get it. This is racism. These are microaggressions.”*
>
> *It’s the society we live in now. You know, in the 70s and 80s, you were supposed to erase [the background of the adoptive child]. That was what was best for the children. And nowadays, everything should be out in the open, because now that’s what’s best for the children.*
>
> *And when these [adoption-related] issues have been given more attention, it’s easier to find people today who… A lot of people have some type of knowledge about these things. For the first time, you’re not saying anything controversial.*
>
> *Society has changed—people talk more about mental health, young people today might say “I have ADHD” or “I have autistic traits” and it’s nothing weird about it. […] And there’s social media and things like that, so I think it’s easier to be open about it today.*

### 3.3 Needs and suggestions

#### 3.3.1 Therapist identity

Regarding preferred therapist characteristics, many participants say that it may be easier for them to open up and address important adoption-related issues if the therapist is also an adoptee or a person of color:

> *I’ve felt for a while now that I’d like to see a therapist who’s also adopted. Because there are so many subtle things that are hard to explain, but for another adoptee it’s like: “Right, you mean that thing”. You might talk in abstract terms, but when you speak to another adoptee, they get it right away. On a much deeper level, without you having to explain. Then of course, you might still misunderstand each other.*
>
> *[S]omeone with a similar background. It’s not that I necessarily have to see someone from Chile, or that a Korean person has to see someone from Korea. I mean, it’s not like that. It might just as well be a native Swedish adoptee. I’m looking for someone who can relate to others through their own life experiences. Otherwise, I reckon they won’t be able to understand, and then they can’t really help either.*
>
> *Perhaps I won’t need to explain that there’s adoption-specific discrimination to someone who’s adopted, you know. Or I won’t need to explain being discriminated against in a certain way, being an Asian woman, to a woman of color. Because, of course, you get tired of explaining yourself all the time. Explaining who you are, or where you’re really from, or why that is. I mean, this constant explaining wears you down.*
>
> *If I get to choose… Maybe it’s a bit more difficult seeing a typical Swede. I guess I’ve instinctively felt—and perhaps I’m prejudiced—but I’ve felt instinctively that: “You won’t understand, so why talk about it?”*

On the contrary, many participants also highlight other aspects of the therapist-patient relationship as more important:

> *I’d say it’s more about chemistry for me.*
>
> *I actually don’t know. When it’s someone professional, I think that person—let’s say it’s someone who specializes in adoptees, I’d expect that person to have adequate training to be able to confront it.*
>
> *Maybe I would feel that there was an expectation that we should click just because we’re adopted. And perhaps we wouldn’t even like [each other]. I think it’s really more about the knowledge of the therapist.*

#### 3.3.2 Designated resource center

When asked about their thoughts on a suggested designated resource center focused on adoption-related issues, including health and well-being, most participants welcome such an initiative:

> *That would be the best choice I suppose, to have everything under the same roof. That there was a center for everything related to adoptees. That’s what I would have wanted when I was younger.*
>
> *A center or a specialist unit. So that everyone knows what to do whenever there’s a problem. Like, “Oh, we have a patient with this problem?”. Then they’d know that “alright, this is where we’ll refer you”. So that you’d never have to face that: “God, I don’t know what to do with you”. Because you’ve had that experience so many times as an adoptee, so many times. There should be a system, a protocol for how to deal with this.*
>
> *If everything was in the same place and they followed you throughout your—sort of like, “Well, you can always come to us with any questions you might have, if you need support or if you need help with seeking your roots or pretty much anything”.*
>
> *It’s probably beneficial, absolutely. I’m thinking that they’ll also collect a lot of information from various cases that can contribute [in building knowledge].*
>
> *Like a community center rather than a healthcare center. They could very well offer healthcare, but it doesn’t need to be like… I’m thinking that it doesn’t necessarily need to smell like in a hospital and have a certain type of art on the walls and people are walking around in these plastic slippers. It could be like… There could be room for things that are… Talking about existential questions or more like… Debates and a little bit of everything.*

Even so, there may also be barriers to successful implementation that need to be considered:

> *It’s a great idea for people living in Stockholm [i.e., the capital of Sweden], but for those living in [the North] or in [the South], they can’t travel to Stockholm just for this. There needs to be a widespread competence.*
>
> *The question is, how do you figure out which adoptees need support from a specialist unit and who can be seen in primary care or by whoever, basically?*
>
> *I can imagine that a part of me would have been provoked, since I’ve had a hard time accepting my Indian background. I wanted to be Swedish. So maybe I would’ve felt upset if someone said: “Come to this place where there are lots of other adoptees and talk!”*

Ensuring easy access to information about currently available resources is more of a priority for some participants, regardless of the format in which it is provided:

> *I would have wanted something—you know: an easily accessible overview of which therapists, what kind of support you can access… Not spread out in 40 different places and you need to look for it. And I’m thinking that I work in communication, it’s part of my job, and if I sometimes find it difficult to locate this information, other people must have a really hard time.*
>
> *The most important thing would’ve been information: “This is where to turn.” […] So you actually know who to contact. If it’s a special center or not, that doesn’t really matter. It’s usually not like that. If you need treatment for cancer, you go to one place [in the hospital] and if you need a speech therapist, you go to another place in another building and so on. So that aspect isn’t important, but it should still be like an umbrella of services: here you can find the information and here you can get further help. That’s paramount, of course.*

#### 3.3.3 Improved knowledge

In response to experiences of limited insight into adoption-related issues among therapists and other healthcare staff, participants call for improved knowledge in several areas, such as adoption trauma and the impact of racism and other post-adoption factors on health:

> *The most important thing is that the therapist is responsive and flexible, and that they’re transparent about what they know and what they don’t know. And that they don’t generalize, since every adoptee has their own experiences and views that need to be respected. That said, of course you hope that the therapist is as knowledgeable as possible, so that they understand the complexity of the adoptee experience.*
>
> *Sure, you could work on increasing awareness in healthcare staff… Like in primary care. It could be more integrated—and I realize it’s a huge apparatus and super difficult—but you could integrate it in medical school, in nursing school.*
>
> *Because ultimately, after all these years I realize that if that individual is trained in White psychology—and I have no idea what they learn about adoption—then I’m not so sure. […] It’s not just about [who you are], it’s about what you’re teaching these therapists, what sort of knowledge?*
>
> *I don’t know if it’s well-known in Sweden, adoption trauma. I’ve never found… I’ve looked for it a lot. It’s big in the United States, for example. They look at it differently, there’s nothing strange about being affected throughout life by what happened to you as an infant. And here, it’s as if the idea that it might affect you today is just so odd.*
>
> *Just as if I’m undergoing surgery, I’d prefer if it wasn’t done by somebody who only performs that procedure once a year. I’d prefer if it was done by a surgeon who had some continuity. Because it builds experience. And I if there are people who’d like to train and specialize in adoption, it would be nice if we were offered that kind of support.*
>
> *Spontaneously, I think it’s important to realize that being an adoptee is a risk factor. […] I know that I may seem as if I’m high-functioning. I have a job and all of that. But that’s also part of being an adoptee. To be like a chameleon. And being a survivor of early trauma—knowing what that does to a person. It’s about survival. So then you survive. I mean, you don’t just lie down and “nah, I can’t do this”. It’s more like… I think I would’ve needed someone to see through me.*

#### 3.3.4 More than cognitive-behavioral therapy

Several participants have experienced that cognitive-behavioral therapy (CBT) has not been sufficient to fully address adoption-related themes and express a need for complementary psychotherapeutic approaches. Of course, CBT comes in many forms and can very well include a focus on adverse childhood experiences, racism, etc.; even so, participants describe how simplistic “here and now” variants of CBT have not always been very helpful:

> *I don’t know, it felt like pure CBT for problems like these was more like: “Your way of thinking is flawed, just learn to think in the right way and it’ll be solved”. I’m thinking that it’s not simply a conditioned phobia, there are other things that need to be dealt with.*
>
> *How would ten sessions of CBT be enough for a lifelong trauma? It just doesn’t add up.*
>
> *Healthcare must be trauma informed. […W]hen it comes to relational trauma, there needs to be “permission, permission, permission” in relation to the client, since permission is exactly what’s been missing and what caused the trauma. I believe this is one of the necessary cornerstones for adoptees to dare to engage and stay in therapy. But I also believe that this is really hard to achieve through therapy in which the therapist is seen as an expert—CBT or psychodynamic [therapy], for example—who’s supposed to evaluate and lead.*
>
> *[I]t’s important to offer a broad spectrum of interventions and treatments. Some adoptees haven’t found their way out of the adoption fog, whereas others have. Some don’t need trauma-focused treatment; others have a great need. Even if you don’t need or don’t take an interest in trauma-focused treatment, you may need help in building a secure attachment within yourself and in your relationships, learning relational skills, [or] developing your mentalizing capacity.*

#### 3.3.5 Applying a life-stage view

Many participants note that when adoption has been addressed in healthcare, the focus has typically been on adoptees as children (which has contributed to the tendency for infantilization described above). However, as adoptees grow older, there is inevitably a need to apply a life-stage view on adoptee health:

> *In various phases in life and at various life events, the need for support may be greater: pregnancy, parenthood, childlessness, romantic relationships, loneliness because you don’t have a romantic relationship, return journeys, reconnecting with your bio family, coming out of the adoption fog, breaking up [or] regaining contact with your adoptive parents, bio parents or adoptive parents passing away, etc.*
>
> *[I]t needs to be a long-term contact, because you’re an adoptee all your life and there will be ups and downs. It’s not like you have ten sessions with somebody and then you’re done […]. Throughout my lifetime, I will face different issues that are connected to my adoption and well-being—I don’t imagine that you’re ever finished with it.*
>
> *Perhaps there should be something for you when you’re in your 20s—like, 23 or 25… When you’re starting to mature a bit and you’re perhaps slightly more susceptible than during your teenage years. And it’s still fairly early in life, early enough so that many people haven’t started a family of their own. The more healed you are before you enter that stage, the better.*
>
> *[B]eing a parent as an adoptee, where are those discussions? And where are the discussions about our White parents passing away, and now we’re the oldest generation here? Nobody talks about that.*
>
> *When you’re 40+ and you’ve gone through all these major things, then you can suddenly focus on yourself. [… You] have healthy children, you have a good family or something, and suddenly, you start to focus on finding yourself again. And then this adoption question arises, as it did for me.*

#### 3.3.6 Education and support for adoptive parents

Many participants mention that adoptive parents need more assistance to be better prepared for potential challenges, such as navigating racial differences and standing up to racism:

> *But then when they receive the child, it’s like: “Ok, bye”. They could use a course on the kind of problems their child might face. Because all adopted children have been through at least one separation. Whether it was brutal or not, there has always been a separation. And I think many parents don’t realize that. That’s what I’ve experienced and what my friends have experienced. It creates a lot of friction between children and parents.*
>
> *If you’re a racialized child in a White society, things will happen. It’s not that your child might experience racism—your child will experience racism. And then you need to be able to help them deal with it.*
>
> *I think my parents would’ve needed… They received no help whatsoever, no training. But I think that if they’d been given the opportunity, they surely would’ve felt better talking to others.*

#### 3.3.7 Follow-up during childhood and adolescence

Just as adoptive parents need support, many participants discuss how adoptees would benefit from some type of structured follow-up during childhood and adolescence:

> *If I were an adoptive parent or worked in the field, I’d most certainly think that there should be some kind of support [for the adoptive child], and for many years. Not just: “Bye, have a good life, we’ll never see you again”.*
>
> *I would’ve wanted some check-ups in my early teens because I’ve understood that’s a tough period for many.*
>
> *But if you’re going to import a lot of children, perhaps you ought to follow up and see that they’re doing well. And not just assume that because it’s an affluent Swedish family, everything will be hunky-dory. It’s like they leave you on your own, in a way.*

Several participants acknowledge that if they had been offered adoption-specific follow-ups during adolescence, they might very well have declined. Knowing that the opportunity existed could nevertheless have been helpful:

> *[M]any adoptees don’t really ask for help, so it’s better to offer it continuously and then you can say no, rather than having to ask and not receive the support you need.*
>
> *Say that when I turn 15, I get a letter to my home: “Hi, we notice that you’re 15 in two weeks. When you’re 15, there will be an opportunity for counseling, if you have questions about your background or your relationships or if you’re dealing with identity issues, for example. Call this number if you ever need to talk. You have ten free sessions.” As an example. Then I would’ve carried that with me, and I would’ve thought about it—it might have taken a few years, but I would’ve carried it with me, and I could’ve contacted them when I was ready.*

#### 3.3.8 Support groups

Several participants mention that it could be useful to meet other adoptees in organized support groups, as a forum for sharing experiences and seeing yourself in others:

> *I think group talks are great. Groups that aren’t necessarily about sad stuff, but also groups that focus on existential loneliness in adoption. I think group sessions like those are great.*
>
> *That’s also very common: “Everybody think I suck, and I agree.” Those type of [ideas] are pretty common for adoptees. So it’s a little bit of—well, strengthening the self-esteem of those people.*

#### 3.3.9 Addressing adoption experiences during pregnancy and as a new parent

In parallel to the somewhat broader subtheme about the need for applying a life-stage view described above, many participants talk about how pregnancy, giving birth, and becoming a parent can be particularly shattering life events for adoptees:

> *I could’ve used more support from my health center. To talk to a midwife about how I felt. What I realized was that I’ve actually been taken from someone, I’ve been in their womb for nine months and then taken, and I felt so very sad. And it was also the first time ever that I felt like—god, I’ve never reflected upon this before—that there was another person who looked like me.*
>
> *[E]specially when you’re pregnant as a woman, it’s incredibly triggering.*
>
> *It can be hard for an adoptee, not least when the kids are the same age as when you were abandoned. It can be a tough period. It becomes so tangible, perhaps, what actually happened.*
>
> *[I]t’s still one of my fears, to not be able to stand up for [my daughter]. If she experiences racism, how will I who never learned to stand up for myself—how can I help her? Or be able to support her? It haunts me as a parent.*

#### 3.3.10 Access to medical check-ups, blood tests, and genetic counseling

Many participants discuss the fact that transnational adoptees typically do not have any—or occasionally very sparse—information on biologically inheritable vulnerability and disease. However, when they mention this to their physicians, it does not necessarily prompt an extended assessment but is more often treated as an anamnestic obstacle that simply cannot be overcome:

> *And now when I’m getting older, there may be lots of inheritable disorders and weird stuff that nobody knows about. And it actually annoys me that I don’t get to do extra blood tests or something like that. Because I don’t know anything. I can’t just go home and ask like most people.*
>
> *Mostly I’m tired of being asked when nobody cares about the answers. They ask because they’re supposed to, and then: “Uh-huh, ok.” I was at a medical check-up the other day and they asked: “So where are you from?” “Well, I’m adopted from Korea.” “Ok, do you know anything about your parents? About their health? Your biological parents?” “Yes, my dad died in a workplace accident and my mom had heart problems and a stroke.” “Ok”, and then they just move on. There’s no… Perhaps they’d want to run an ECG? See what I mean? Me telling them this doesn’t lead to anything.*
>
> *When I was pregnant, they asked: “What was it like when you were born?” I have no clue. No? So then maybe they should do something with [that information] or at least comment on it, like “alright, that’s a pity, then we’ll need to keep a few doors open and follow up on it”. It doesn’t need to be any more difficult.*

#### 3.3.11 Better knowledge about medical conditions that more often affect persons with a non-European background

Several participants also express a need for improved knowledge in healthcare about medical conditions that more often affect persons with a non-European background, including competency in the assessment of melanin-rich skin:

> *I was in the school nurse’s office all the time because of abdominal pain. They never figured it out. And now that I’m grown up—I’m lactose intolerant. I can’t drink milk and that starts when you’re six or so, I guess, but nobody in [small Swedish town] thought about that. And it’s not exactly rocket science today.*
>
> *And apparently, it’s very common in China and other Asian countries. And I had to google it to find out. I showed it to a doctor several times and their response was just: “Nah, it’s probably nothing”. […] All this extra work you need to do if you’re not part of the norm, that’s just how it is.*
>
> *I was there because of this [rash]—and this person, she said that she couldn’t see it on my skin because they don’t learn that, apparently. To see it on darker skin. You need to have light skin.*
>
> *Eventually, I had an emergency delivery, and he came out weighing 5600 grams [i.e., 12.5 pounds]. […] And I thought: What if they had been able to find a weight and length chart for pregnant Vietnamese women and compare it to a Swedish chart? Would they have made different decisions based on that?*

Of course, some of these problems affect other individuals with a non-European background in Sweden too; however, the fact that transnational adoptees often do not have access to a community of people from the same part of the world to turn to for basic medical advice, makes it more of an obstacle.

#### 3.3.12 Economic resources

Because of the steep financial costs described above associated with private psychotherapy, return journeys, and seeking one’s roots, many participants call for access to economic resources as part of an effort to improve adoptee health:

> *You shouldn’t have to use your own savings for this. And it shouldn’t become a class thing. Not only those who can afford it should have access to adequate help.*
>
> *This type of post-adoption service must be free of charge, of course. The government just needs to step up and finance it. We as adoptees can’t be expected to pay for this ourselves.*
>
> *I reckon it would have to be a standard amount. It could go through the Social Security Agency, and you’d log on and: “Hello, you have this amount that you can use. These services are available.” And then you could see [therapists] who are affiliated.*

Several participants also talk about the fact that adoptive parents receive financial support when they adopt a child, whereas adoptees do not have access to similar support, as a fundamental injustice:

> *[L]ooking at the budget and the subsidies that have gone to adoptive parents so they can adopt a child—it’s a lot of money. And if we now know that adoptees often need to seek help, but don’t receive any economic support or subsidies, I think we often feel that it’s unfair. And it’s a signal about what’s prioritized.*

#### 3.3.13 Support for children of adoptees

Finally, many participants point to children of adoptees as a group in need of more attention in healthcare. Even though this is a heterogeneous population, children of adoptees all in some sense belong to a ‘second generation’ that shares background and experiences with the adoptee parent:

> *This is a multigenerational trauma. My children don’t have a clue either about their family and relatives and culture and all that. The just know that something bad happened and now all those other things are gone. They miss it too. Also, my children—my daughter more than my son perhaps, but I think my son has also been racialized and experienced racism. My daughter even more than I did when I was [little].*
>
> *It can also be helpful to be aware of it as a risk factor among second generation adoptees. That they too can feel uprooted, not having any contact with their biological family. That they can inherit this rootlessness. It’s an interesting time now, I think, because the children of adoptees are becoming old enough to reflect upon their lives on their own, and perhaps become parents themselves.*
>
> *Adoption doesn’t end with the adoptee. I’m not saying that there need to be things to deal with or problems, but I’m thinking there’s still a risk. Just the fact that their parent may not feel so good all the time.*

Notably, the lack of information about biological heritability is also a factor for children of adoptees:

> *[T]he doctors ask: “Do you have heart disease in your family? Diabetes? Any other diseases run in your family?” That triggers me so much I feel like throwing up, every time. I don’t know. I’ll never know because some motherfucker stole me. And my children don’t know either. It’s inherited, unfortunately.*

## 4 Discussion

The findings reported here, based on individual in-depth interviews with 65 adult transnational adoptees in Sweden and written answers from one additional participant, point to a large number of adoption-related issues that are of immediate relevance to healthcare and that underscore the need to further attend to the health and well-being of transnational adoptees. Participants describe several barriers in accessing adequate treatment. Some of these, such as a lack of insight into and interest in adoptee health, are shortcomings directly attributable to healthcare. Other barriers involve healthcare but are also characteristic of society at large and may contribute to feelings of resignation, lack of trust, and a reluctance to even seek support and treatment for fear of being invalidated and mistreated; these include colorblindness and unwillingness to address racism, expectations of gratitude, disregard for personal boundaries, and tendencies of infantilization of adult adoptees. Yet other barriers exist in the form of steep financial costs, lack of support from adoptive parents, and a fundamental mistrust of support structures that directly involve adoptive parents, adoption organizations, and adoption-related authorities. Participants also describe resources that are helpful in dealing with health-related issues, such as the community of fellow transnational adoptees and the recent media exposure of illegal adoptions.

In response to the many identified barriers, the study participants discuss health-related needs and suggestions for the development of adequate support for transnational adoptees. These include more well-defined and easily accessible structures of support, improved knowledge and competence in healthcare, a broader psychotherapeutic repertoire that better addresses adoption-related themes, routine follow-up during childhood and adolescence, education and support targeting adoptive parents, and economic resources. Moreover, participants underscore the need for adequate support in situations that may be particularly stressful for transnational adoptees, such as during pregnancy and as new parents. They point to the importance of being offered medical check-ups, genetic counseling, etc. to compensate for the fact that most transnational adoptees do not have any information on heritable diseases that may run in the family, and improving the knowledge on medical conditions that more often affect persons with a non-European background. The need for greater attention to the well-being of children of transnational adoptees is also highlighted.

Themes such as experiences of racism or disregard for personal boundaries among transnational adoptees have certainly been raised before in the research literature, in a Swedish context as well as elsewhere (29,30,37,38). Importantly, the present study demonstrates how these experiences also affect and shape adoptee health, encounters in healthcare, and help-seeking patterns. For example, an omnipresent societal narrative of gratitude—i.e., a view of transnational adoption as morally righteous (44), accompanied by expectations that adoptees ought to feel lucky for having been “saved”—does not only create ambivalence in terms of personal identity, but also makes it substantially harder for many adoptees to seek support and treatment in times of need, out of fear that they will upset their adoptive parents or be invalidated and dismissed by therapists. Likewise, experiences of ‘othering’, racism, and colorblindness may have a direct impact on trust and help-seeking. Study participants describe a general unwillingness among therapists to acknowledge and discuss experiences of racism, a tendency that is clearly associated with a prevailing idea of Sweden as a ‘post-racial’ society in which a comprehension of race as an outmoded and unscientific concept dominates in clinical settings (45). To be clear, we see race as a social construct that is nevertheless ‘real’ insofar as notions about race and racialization have a profound impact on people’s lives. Moreover, we adhere to a view of ‘racisms’ as a plurality, acknowledging that whereas there are certainly elements that are common to various forms of racist discourse, there are also particular traits that shape anti-Black racism, anti-Asian racism, antisemitism, etc. A sensitivity to the specific forms or facets of racism that affect the individual patient (46) and an openness to explore intersections of race, gender, socioeconomic status etc. (47) are therefore crucial in psychotherapy with transracial adoptees. Not least, basic psychotherapeutic components such as the ability to simply listen, empathize, and validate must take center stage in dealing with experiences of racism, just as with any other patient experiences (48); our participant narratives clearly demonstrate that this is rarely the case in a Swedish ‘post-racial’ context. Moreover, therapists must be open to supporting patients in tasks such as resisting internalized racism and self-blame, navigating supposedly colorblind attitudes in society and adoptive families, and envisioning healing (30,49). A general hesitancy to address these aspects of patients’ lived experiences inevitably makes therapy less relevant for many groups. Importantly, there is also a very real risk of encountering racism in healthcare settings; this is well-documented in Sweden as well as in other parts of the world (50).

The reported unwillingness among therapists and healthcare staff to address adoption-related issues may perhaps surprise readers who are accustomed to a view of therapists as characterized by a general openness to the exploration of the most diverse—and extreme—life experiences of their patients/clients. We can only speculate in the reasons behind this apparent blind spot among many Swedish therapists. Several participants describe how they have encountered the gratitude narrative discussed above in direct contacts with healthcare staff; if therapists are themselves heavily influenced by prevailing notions of transnational adoption as primarily “good” or “beautiful”, this may obscure alternative narratives and make it difficult to empathize with adoptee patients who are perceived as engaging in unprovoked “self-pitying”. Moreover, it has been suggested that one of the performative functions of the adoptive family within a Swedish civic discourse has been to absorb Otherness and uphold a fundamental notion of Swedishness in which race and adoption background are seen as irrelevant (51). If it is difficult for many therapists to deal with patient experiences of racism in a ‘post-racial’ setting, it may be even harder to relate to the specific experiences of ethnically ambiguous ‘in-betweenness’ of transnational adoptees in this context. There is an obvious need for improved knowledge about health consequences of transnational adoption, as well as strengthened structural competency (52) related to issues such as racism and trauma. Moreover, calls for the decolonization of mental health and psychotherapy through, for example, increased attention to hidden biases and stronger emphasis on racial awareness (53–55) could be further incorporated into the medical curriculum and professional competence training (56,57). Our study participants are somewhat divided on the significance of therapist identity. Some express that it is easier for them to discuss their experiences as transnational adoptees with an adoptee therapist or a therapist of color, whereas others put greater emphasis on personal chemistry or general therapeutic skill. Even so, many of those who prioritize aspects unrelated to therapist identity also say that—everything else equal—a therapist of color may be preferable. Meeting patient preferences regarding patient-therapist matching can probably contribute to an increased likelihood of opening up and daring to approach sensitive topics in treatment (58). Addressing a general lack of diversity among therapists should certainly be a priority; nevertheless, there is undoubtedly a parallel need for improved awareness of and competency in dealing with issues relating to transnational adoption among White therapists.

It is also worth mentioning the role of the transnational adoptee in performing so-called affective labor—a labor that, in this case, involves the fulfillment of family dreams and expectations of love and attachment (24) as well as the affirmation of a national self-image of righteousness and colorblindness (44). Returning to the breaches of integrity discussed above, participant descriptions of sudden outbursts of sentimentality among otherwise professional therapists when the topic of adoption is brought up point to the adoptee as a projection screen for various wishes, hopes, and expectations in society. Since no one is immune to culturally ubiquitous images and stereotypes, therapists must learn to actively identify and temper their own preconceived notions of adoption in treatment contacts with transnational adoptees, so as not to risk alienating and harming patients/clients.

At the risk of stating the obvious, many participants express great affection toward their adoptive parents while simultaneously acknowledging experiences of having been let down or not fully supported at decisive moments. There is obviously substantial skepticism toward the involvement of adoptive parents or adoptive organizations in counseling and treatment. In light of recent media reports of far-reaching illicit adoption-related activities, including outright trafficking of children from the Global South (6,7), it is not surprising that many transnational adoptees currently lack fundamental trust in adoption organizations and adoption-related authorities. This lack of trust also affects healthcare, not least in a largely government-run public healthcare system.

Study participants are generally positive toward establishing a national research and knowledge hub on transnational adoption, as suggested by a previous official report of the Swedish government (36). From a strict healthcare viewpoint, this type of designated facilities tailored to the needs of specific minority groups in society have traditionally been a rarity in Swedish and European healthcare systems (59), whereas in North America, it is more common to find facilities explicitly addressing the healthcare needs of the Hispanic/Latino or Asian American populations, for example (60). The benefits of designated healthcare facilities or resource centers include opportunities of catering to unmet needs in the population, creating protocols for addressing minority health disparities, and signaling the importance and urgency of improving access for underprivileged groups. However, there are also possible disadvantages of creating “niche” facilities. For example, competence in generalist healthcare may suffer if the creation of designated facilities signals that the topics at hand are beyond the realm of general practitioners. As some of our participants mention, determining which adoptees to refer to a specialist center and which are better served by generalist healthcare is not necessarily a straightforward task. Moreover, facilities targeted to specific groups are more vulnerable to political volatility. All things considered, the findings reported here make us lean toward creating of one or more national research and knowledge hubs on transnational adoption. Hopefully, such hubs could bring together multiple disciplines under the same roof. A recent Dutch governmental inquiry into illicit adoption-related activities (7) recommended the establishment of a National Centre of Expertise integrating knowledge about psychological treatment, legal support, and root-seeking. A similar Swedish center could potentially learn from Dutch experiences.

Last, it should be noted that recent media exposure and formal investigation concerning the widespread occurrence of illegal adoptions (6,7) are mostly seen as positive resources among the study participants. Whereas some participants describe increased anxiety upon learning about these far-reaching illicit activities, most experience the substantial media attention as empowering. This points to official acknowledgment and recognition of wrongdoings as central to adoptee well-being and health.

### 4.1 Strengths and limitations

The present study explores experiences, opinions, and needs concerning healthcare among adult transnational adoptees, a topic that has not been sufficiently addressed in research. For a qualitative study, the number of participants subjected to individual in-depth interviews is relatively large, ensuring a breadth of perspectives. Furthermore, involving a reference group of transnational adoptees in various roles ensures that our research questions have some established resonance among the population under investigation.

The findings presented here should also be interpreted in light of a couple of limitations. Participants were recruited by disseminating information about the study on websites, in newsletters, and on social media accounts of various Swedish organizations for transnational adoptees. This may possibly have favored recruiting of participants who are already knowledgeable about and dedicated to adoptee rights. Of course, this need not be a limitation in a study about the improvement of adoptee health; however, it should be mentioned as a possible bias. Importantly, transnational adoptees join adoptee organizations for a plethora of reasons, including interest in adoption critique and activism, as well as to meet friends, hang out, and have fun. Our impression is that the study participants demonstrate a variety of attitudes toward adoption: some are highly critical of the adoption industry, whereas others are not. Notably, while several participants explicitly state that they do not feel fully comfortable in a context of adoption critique, the narratives of these participants are not fundamentally different from the rest. Those who are hesitant to take an “activist” stance nonetheless describe decisive experiences of racism, colorblindness, expectations of gratitude, etc., and discuss these topics in a detailed and thoughtful manner; thus, although participant accounts vary in terms of attitudes toward the adoptee community as a social and political entity, they do not necessarily differ significantly in content.

Naturally, healthcare systems vary widely across different countries and regions. This study was conducted in Sweden; however, we expect the findings reported here to be of significant relevance to other settings too, given that many of them reflect transnational adoptee experiences and needs of a more fundamental nature that are not necessarily strictly linked to a specific geographical location. Nevertheless, some of the findings and implications reported here may be less applicable in a non-Swedish healthcare context.

### 4.2 Implications for the future

Based on the findings reported here, the following ten recommendations can be made:

1. Measures should be taken to improve knowledge about adoption-related issues and adoptee health among healthcare staff, through the incorporation of relevant aspects into medical curriculums and professional competence training. A systematic approach to the improvement of knowledge about medical conditions that more often affect persons with a non-European background (including competency in the assessment of melanin-rich skin) is needed, so that it does not merely become a matter of individual commitment.
2. Psychotherapeutic competence in addressing issues related to racism should become a priority. This also includes ensuring greater therapist diversity in terms of background and race in public healthcare to meet the needs of those who experience patient-therapist matching as instrumental in being able to open up and address these issues in a helpful way.
3. The current focus on basic CBT in Swedish mental healthcare should be complemented by a broader psychotherapeutic repertoire that better addresses adoption-related themes.
4. One or more national research and knowledge hubs on transnational adoption should be created, possibly based on Dutch experiences. Careful consideration is needed regarding the specific format and purpose, stakeholder involvement, geographical location, procedures for referral, etc.
5. Routine follow-up of transnational adoptees during childhood and adolescence should be initiated. The optimal format of post-adoption services needs further exploration. Mandatory follow-up during adolescence may be alienating and counterproductive for some; a standing offer arrangement is potentially more acceptable.
6. To the extent that transnational adoption to Sweden continues, prospective adoptive parents should receive training and education to be better prepared for potential challenges, such as navigating racial differences and standing up to racism.
7. Healthcare staff involved in antepartum, intrapartum and postpartum care should receive training and education to understand better and meet adoption-specific issues that may arise during pregnancy and as a new parent.
8. In response to a lack of information about inheritable conditions, transnational adoptees should be offered routine medical check-ups, extended blood testing, and genetic counseling.
9. Economic resources should be made available to support transnational adoptees in accessing psychotherapy and treatment (if adequate access cannot be provided through a research and knowledge hub). Moreover, there is an ethical imperative to support adoptees who wish to visit their country of birth or engage in seeking their roots.
10. The needs of children of adoptees—the next generation—should be further addressed in research and treatment.

This is not an exhaustive list; there are of course a multitude of others measures that can be taken on various levels in society to improve adoptee health and well-being. However, based on the interview data reported here, these ten recommendations can be regarded as a starting point in an effort to ensure the provision of and access to adequate prevention, treatment, and follow-up for transnational adoptees.

## 5 Conflict of interest

The authors declare that the research was conducted in the absence of any commercial or financial relationships that could be construed as a potential conflict of interest.

## 6 Author contributions

NH and MS jointly conceived of and designed the study. MS administered the project. NH and MS both performed patient interviews and analyzed the data. MS prepared the original manuscript draft. NH reviewed and edited the manuscript. MS provided oversight and general supervision.

## 7 Funding

NH performed five interviews as part of a research track during her residency program in psychiatry in Region Stockholm. Apart from this, no funding was provided for this study.

## 8 Data availability statement

The full dataset analyzed in this study is not available, for reasons of patient confidentiality. An aggregated and anonymized dataset is available from the corresponding author upon reasonable request.

## 9 Acknowledgements

We would like to extend our sincere thanks to all study participants who bravely shared their personal stories with us. We would also like to express our gratitude to the members of the project reference group—Anna Amazeen, Marit Arnbom, Bonnie Berggren, Anna Jin Hwa Borstam, Tobias Hübinette, and Lisa Wool-Rim Sjöblom—for their generous contributions to the completion of the study.

## Notes

### Competing Interest Statement

The authors have declared no competing interest.

### Author Declarations

The Swedish Ethical Review Authority gave ethical approval for this work (Nos. 2022-03422-01 and 2023-03465-02).

